# Time of Day as an Unmeasured Confounder in Oncology Trials

**DOI:** 10.64898/2026.03.05.26347742

**Authors:** Jonathan Somer, Gal Benor, Ayelet Alpert, Ruth Perets, Shie Mannor

## Abstract

A recent randomized clinical trial in non-small cell lung cancer^1^ confirms what numerous observational studies have reported - time-of-day (ToD) may dramatically influence treatment outcomes in cancer patients^2–9^. In this recent trial median overall survival (OS) decreased from 28 months in the early ToD arm to 16.8 months in the late ToD arm. We raise the concern that clinical trial outcomes may be influenced by seemingly minor biases in treatment time across arms. We also suggest that by measuring or randomizing treatment-time in clinical trials, we may identify beneficial ToD-dependent treatments that would otherwise be overlooked.

## Time of day may influence outcomes of randomized clinical trials

Biasing the time of treatment by a few hours may seem negligible in a large clinical trial with years of follow-up. However, a recent randomized clinical trial in lung cancer displayed a remarkable increase in survival when treatment (immune checkpoint inhibitors together with chemotherapy) was given earlier during the day^10^. The hazard ratio (HR) for survival in this study was 0.42^10^. This HR is consistent with previously reported effect sizes from observational studies^2–8^. For example, a meta-analysis that compared early vs late ToD immunotherapy across five cancer types, estimated a HR of 0.5 (CI: 0.42-0.61)^8^.

This effect size is striking. For comparison, physicians consider HRs of 0.6-0.8 to be clinically meaningful^11^, and the FDA approves approximately half of drugs with a HR of 0.8 and over 90% of drugs with a HR of 0.7^12^.

Due to this large effect size, a relatively small bias in the time of treatment may produce a significant difference between treatment arms. Namely, if physicians prefer to administer the experimental drug earlier during the day, the observed advantage of the new drug may be attributed to this bias. A similar concern was previously raised in the context of CAR-T therapy^7^.

To illustrate this, we simulated a clinical trial comparing two identical drugs but different treatment times per arm (Figure 1). We assumed the effect depends on the ToD based on data by^4^ (Methods). We sampled patient treatment times based on empirical data from two centers^4^. In the control arm we sampled treatment times from the entire day. In the experimental arm we sampled a fraction of treatment times from the first part of the day (Methods). This is sufficient to produce an effect size similar to many approved drugs^12^ (Figure 1).

**Figure 1:**
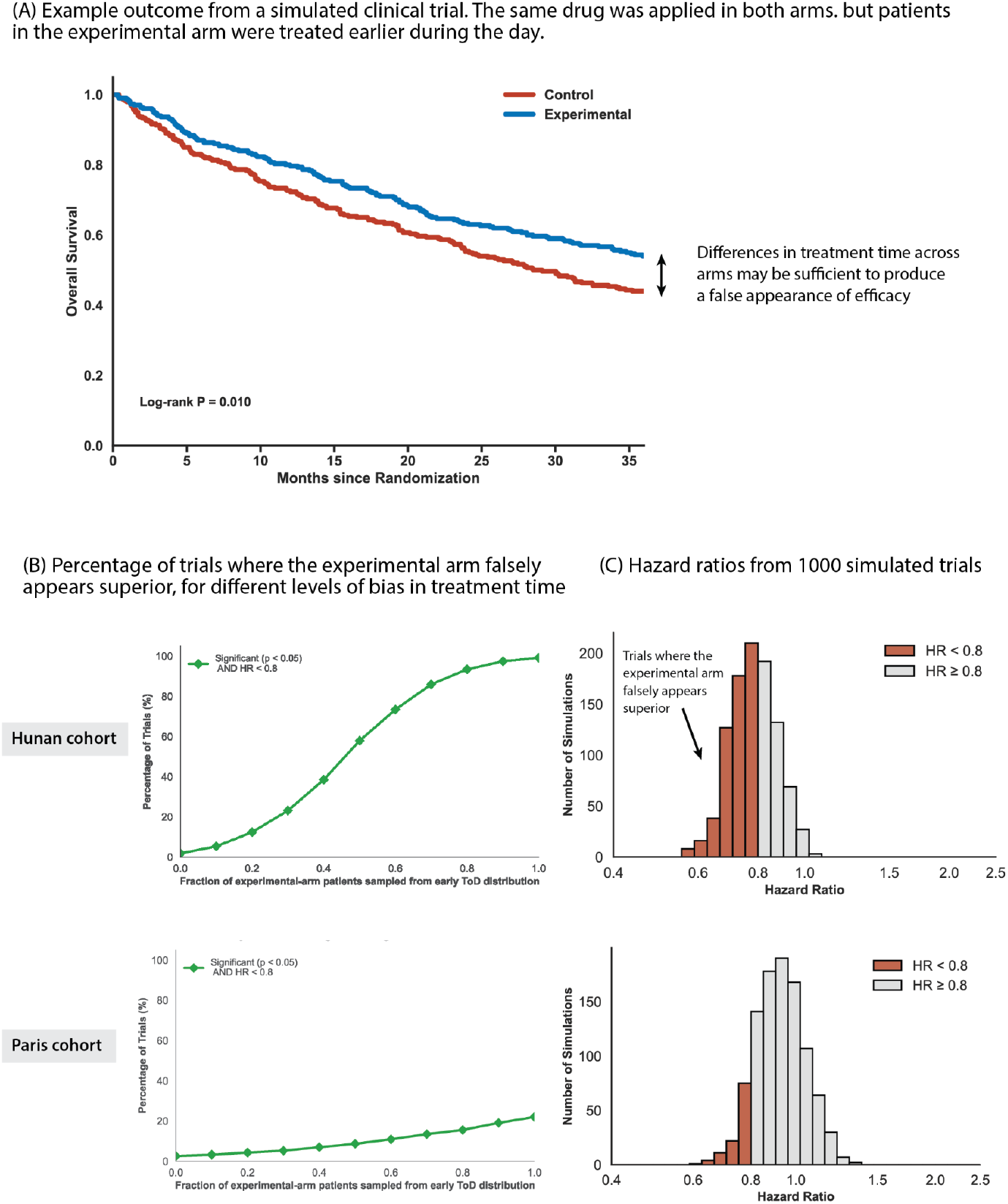
Biased treatment time may influence clinical trial outcomes. We simulated clinical trial data comparing two identical drugs, but different treatment times. The effect in both arms varies throughout the day based on^4^ (Methods, S12C). Patients in the control arm are treated according to empirical treatment times from the entire day^4^ (S12A). In the experimental arm, treatment times are sampled similarly, but a fraction of them is sampled from the first part of the day (from the empirical distribution truncated at the median treatment time, S12B). Panels A,B,C use treatment times from the Hunan, China cohort (S12). Panels D,E use treatment times from the Paris France cohort (S13) A) Example simulated clinical trial outcome. log-rank p-value: 0.0104, HR:0.746. B) Percentage of simulated trials where the experimental arm falsely appears superior, for different fractions of experimental arm treatment times sampled from the early ToD distribution (S6). C) Distribution of the inferred hazard ratio from 1000 simulated clinical trials with 300 patients per arm. Half of experimental arm treatment times were sampled from the early ToD distribution. 58.9% of simulated trials were significant (p<0.05, S3C). 57.7% of simulated trials produced a HR < 0.8 (red). The median HR was 0.7818 (percentiles 2.5 and 97.5: 0.6315, 0.967), median p-value was 0.0278 (percentiles 2.5 and 97.5: 3. 749 * 10^−5^, 0.7518) D) Same as B, for the Paris cohort (S11) E) Same as C for the Paris, France cohort. 8.6% of trials were significant (S9C), and 11.3% had a HR < 0.8. The median HR was 0.9238 (percentiles 2.5 and 97.5: 0.7411, 1.1728) and the median p-value was 0.3942 (percentiles 2.5, 97.5: 0.01485, 0.9668). Supplementary figures 1-11 summarize results from other simulation settings.

We find that biasing treatment time has a larger influence on trial outcomes, in centers with a higher variability in treatment time. In the Hunan, China cohort, treatment times varied considerably throughout the day (mean: 11:43, std: 132 minutes, S10). Biasing treatment time in as few as 20% of experimental arm subjects, produced a false-positive (i.e significant trials with HR<0.8) rate of approximately 12%. When we biased treatment time in half of the experimental arm subjects, the false positive rate increased to 57%. In the Paris, France cohort, treatment times varied less (mean: 10:55, std: 84 minutes, S11). When we simulated trials based on treatment times from this cohort the false positive rate dropped substantially, e.g 8.7% false positives when half of experimental arm treatment times are biased. Supplementary figures 1-11 display results from simulations using a range of settings.

Treatment time may be biased due to a number of reasons. In medical practice it is standard to schedule complicated or risky treatments earlier during the day. This ensures hospital resources and senior staff are available for assistance in case of complications^13^. For similar reasons, physicians may prefer to administer experimental drugs earlier during the day. It is difficult to confirm this bias because the exact time of treatment isn’t frequently logged. Nevertheless, physicians know if a patient was allocated to the experimental arm in up to 88.7% of oncology clinical trials^14^ (open-label studies), making such a bias feasible. This type of bias may also apply to retrospective studies. Other factors that can influence treatment-time may lie outside the control of physicians. These include the overall load on the clinic, delays in lab tests, preparation of the treatment, and of course the time a patient arrives.

As a special case, we note that even if an experimental drug doesn’t have a time-dependent effect, a clinical trial could produce a false appearance of efficacy due to the primary drug. For example, this could occur if the treatment arm receives a combination of a standard immune checkpoint inhibitor (e.g., nivolumab) together with a novel antibody. If the treatment arm is treated earlier during the day, we can expect the enhanced effect of the checkpoint inhibitor to manifest in the overall response rate of the experimental arm. Conversely, if it takes longer to prepare the treatment for the experimental arm, and hence patients are treated later during the day, the reduced effect of the checkpoint inhibitor may mask the benefit of the novel combination.

## Clinical trials should evaluate time-dependent treatment effects

In the study by^1^ modifying treatment time produced an effect similar to the landmark studies that introduced immune checkpoint inhibitors to lung cancer in the first place. For example, in KEYNOTE-189, a trial evaluating the role of the addition of immunotherapy to chemotherapy in treatment naive non-small cell lung cancer patients, the hazard ratio for death was 0.49 (95% CI, 0.38 to 0.64^15^), and in KEYNOTE-024, a trial comparing immunotherapy alone against chemotherapy, it was 0.62 (95% CI, 0.48 to 0.81^16^) The remarkable difference in treatment-effect between morning and afternoon may be leveraged to increase the success rate of clinical trials.

Figure 2 shows results from a simulated clinical trial with a factorial 2×2 design where treatment-time is randomized in addition to the treatment arm. We simulated two drugs with similar average effects, but different time-dependence. The effect of drug A decreases throughout the day, whereas the effect of drug B is fixed. Treatment times were sampled randomly from early and late ToD in both arms. A standard 2-arm trial misses the advantage of Drug A in the morning (Figure 2, left). A factorial design reveals that application of Drug A in the morning outperforms the control (Figure 2, right). A suitable power analysis should be conducted to test if a factorial design is suitable for each trial. Generally, we find that a factorial design may outperform the standard approach when the effect of treatment-time is stronger than the effect across treatment arms (S15, Methods).

**Figure 2:**
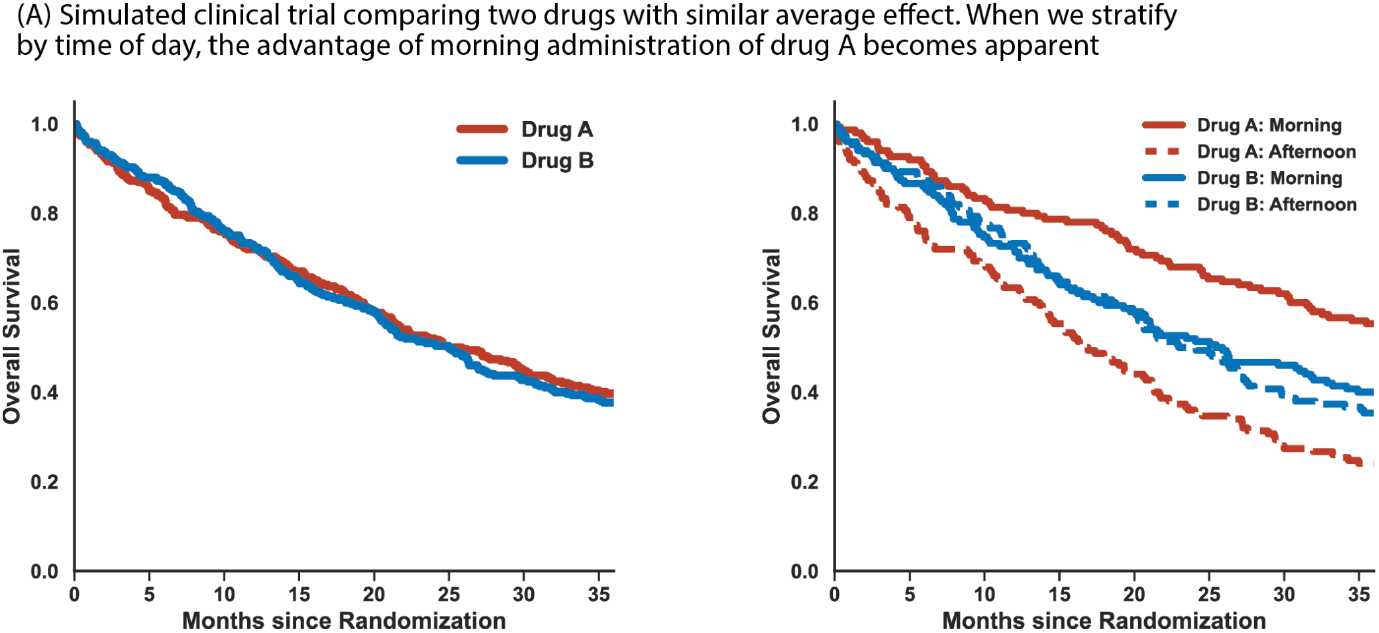
Randomizing treatment time in clinical trials can reveal promising drugs. A) Simulated clinical trial comparing two drugs with similar average effect (Methods). The effect of drug B is constant throughout the day (hazard based on typical median survival in lung cancer, Methods). Drug A has a time-dependent effect - it is better than drug B in the morning (HR=0.6) and worse in the afternoon (HR=1.4). The HRs of drug A were selected such that the two groups display a similar effect without stratification (left, p-value=0.69). When we stratify by time of day, the advantage of morning administration of drug A becomes apparent (right). We used n=300 patients per arm and a follow-up time of 3 years. P-values were computed by pairwise log-rank tests (Bonferroni corrected in the stratified analysis). All comparisons between the morning version of Drug A and other arms were significant (all Bonferroni-corrected p-values < 0.05, S14).

Time-optimized therapies may be challenging to implement in practice. It could be difficult for medical personnel to simultaneously attend to many patients, and patient needs may conflict with optimal timing. Nevertheless, it is important that clinicians are aware of the optimal timing windows for drugs, and a reasonable effort should be applied to treat patients within this window.

Circadian rhythms govern numerous physiological functions, including immune-cell trafficking between lymph, blood, and tissue^17^. Hormones and neural pathways regulate these rhythms and may serve as biomarkers for personalizing treatment time^18^. Furthermore, understanding the physiological mechanisms underlying these time-dependent effects may inspire interventions that mimic the optimal timing, such as drug-hormone combinations, or interventions on lifestyle components such as sleep, exercise, or nutrition.

## Summary

We raise the possibility of a bias in clinical trials due to time-dependent response to treatment. Currently, time-of-treatment is often an unmeasured confounder, which may be present despite randomization. To quantify the extent of this bias and improve our estimates of drug efficacy, we have to measure the time of treatment in future trials and stratify it properly. Reporting guidelines such as CONSORT should require future trials to report the time of treatment. Logging treatment-time is relatively simple to implement, and the additional costs for trials are negligible. Furthermore, randomizing treatment time in clinical trials may reveal promising and harmful drug-timing combinations that would otherwise be overlooked.

## Data Availability

All data produced in the present work are contained in the manuscript

## Methods

### Clinical trial simulation

To simulate a clinical trial we perform the following steps for each patient. We first sample a treatment time. We then sample the event time (e.g death) from a distribution with a hazard corresponding to the sampled treatment time. Prior to simulation we defined a follow-up period of 3 years. Event times that passed the follow-up-period were truncated and censored.

### Sampling treatment times

In Figure 1 we sampled treatment times based on published treatment-time distributions from two cohorts by^4^. We refer to the cohorts as the Paris (France) and Hunan (China) cohorts. We extracted the treatment time data from images of the published figures by^4^ (S12A, S13A; we used cv2 package, version 4.13.0). For the control group, we sampled treatment times directly from the empirical distributions. To bias treatment times towards the early ToD in the experimental group we specified a cutoff between early and late ToD and sampled treatment times from the empirical distribution up to that hour. Main figures display results for a cutoff based on the median treatment time in each cohort (12:00 and 10:30 for the Hunan and Paris cohorts respectively). S1,7 show that similar results are obtained for other cutoffs. Supplementary figures 3-6 and 9-11 show how the inferred effect size changes as a function of the fraction of patients in the experimental arm sampled from the early ToD distribution.

### Sampling death event times

We sample death event times using Weibull distributions^19^. A Weibull distribution is defined by two parameters: λ,*k*. The parameter *k* controls the change of hazard over time. For *k* < 1 the hazard decreases with time; *k*= 1 implies a constant hazard (in this case, the survival function is exponential with a hazard of 1/λ), and for *k*> 1 the hazard increases over time.

We infer the parameters of the Weibull distribution for each hour of the day in two steps. First, we estimate the rate parameter λ for patients treated at 13:00, denoted as λ_13:00_. For a given *k*the parameter λ can be derived from the median survival time. The survival function is given by 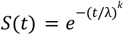. At the median survival time *M* we have: 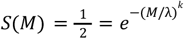 and hence 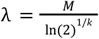. We use this formula to compute λ_13:00_. We assume *M* = 25. 5 months (median for the entire cohort by ^4^ and similar to the 26.3 month median OS from KEYNOTE-02^16^). Figures S1,7 show that similar results are obtained for other values of *M*.

We then infer the hazards for other time points using published hazard ratios from^4^, which compare the hazard in each ToD against the hazard at 13:00 (S12C; extracted from^4^ using cv2 package, version 4.13.0,). The Weibull hazard function is given by 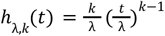 For a fixed *k*, the hazard ratio is constant over time (even though the hazards may change): 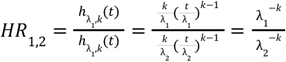. Thus, for each hour, e.g 11:00, we compute the corresponding rate parameter as: 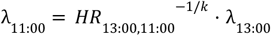

The computations above assume *k*is known. S1 shows similar results are obtained for a wide range of *k*, from 0.5 to 2 (S1). In the main figures we used *k*= 1 (constant hazard).

### Comparing Statistical Power of Standard vs. Factorial Trial Designs

Figure 2 presents a simulated trial where a 2-arm design fails to detect the advantage of the morning version of treatment A. Here we compare the statistical power of a standard 2-arm design against a factorial 2×2 design that tests for time-dependent effects.

We compute the statistical power of both approaches using simulated trials. We first specify the hazards for each of the four arms (the *H*_1_ hypothesis). We then simulate event times for each of the arms based on the chosen hazards. The two-arm trial succeeds if there’s a significant difference between events from A vs. B. The four arm trial succeeds if the superior version of the experimental treatment outperforms all other arms (i.e both versions of the control as well as the inferior version of the experimental treatment). We use a log-rank test for all comparisons with α = 0. 05. Finally, the statistical power is the fraction of successful simulations under each approach.

S15 shows how the statistical power of the two designs is influenced by our assumptions on the main effect (drug A vs. drug B) and time-dependent effects for each drug. As before, we use Weibull distributions with *k*= 1 and a follow-up period of 3 years.

To specify the four hazards, we first assume the hazard ratios for the time-dependent effect in each arm, 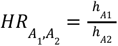 and 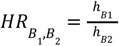. Specifying the main effect isn’t as straightforward because the hazard of a mixture of two populations changes over time (because the patients with a higher hazard are lost more rapidly). We assume that at the end of the trial, survival in the control arm matches the survival in a cohort with a fixed hazard *h*_*B*_ and a median OS of *M* = 25. 5 months (median of the entire cohort by^4^). Namely, 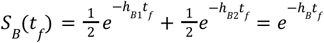. We then specify *HR*_A,B_, the effect size across arms and use it to compute an aggregate hazard for the treatment arm via *h*_A_ = *HR*_*A*,B_ · *h*_B_. As with the control arm, we assume that overall survival in the treatment arm matches that of the fixed hazard at the end of the trial 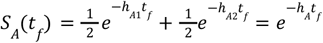. These assumptions are sufficient to compute the four hazards.

## Supplementary Guide

### Simulations based on treatment times from the Hunan, China cohort

S1 - Inferred hazard ratios for varying simulation parameters

S2 - Fraction of significant trials for varying simulation parameters

S3 - Distribution of HRs and p-values for different fractions of experimental arm patients sampled from the early ToD period

S4 - Example Kaplan Meier curves for different fractions of experimental arm patients sampled from the early ToD period

S5 - Simulations with different fractions of experimental arm subjects sampled earlier.

S6 - Fraction of simulated trials with significant (p<0.05) and/or strong effect (HR < 0.8) for varying fractions of experiment arm patients treated earlier

### Simulations based on treatment times from the Paris, France cohort

S7 - Inferred hazard ratios for varying simulation parameters

S8 - Fraction of significant trials for varying simulation parameters

S9 - Distribution of HRs and p-values for different fractions of experimental arm patients sampled from the early ToD period

S10 - Simulations with different fractions of experimental arm subjects sampled earlier.

S11 - Fraction of simulated trials with significant (p<0.05) and/or strong effect (HR < 0.8) for varying fractions of experiment arm patients treated earlier

### Treatment time and hazard ratio by ToD data used in simulations

S12 - Hunan, China cohort treatment times and time-of-day dependent hazards extracted from Huang, 2025

S13 - Paris, France cohort treatment times extracted from Huang, 2025^4^

### Comparison of two-arm vs four-arm clinical trial design

S14 - Pairwise log-rank comparisons between Drug A and B, morning and afternoon arms

S15 - Difference in statistical power between four-arm and two-arm designs

**Supplementary 1.**
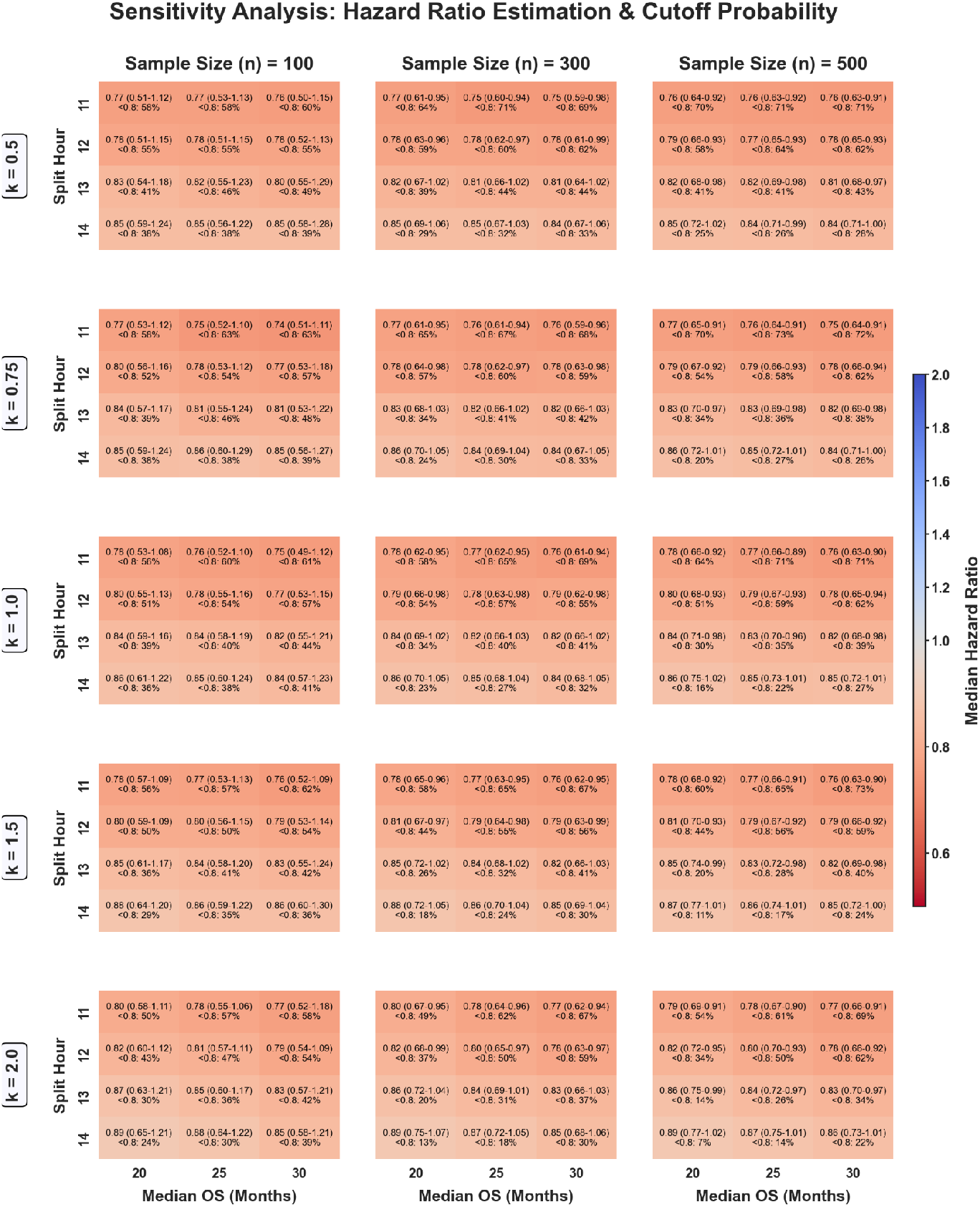
Inferred hazard ratios for varying simulation parameters. Inferred hazard ratios from 1000 simulated trials with 300 subjects per arm. We compare two drugs with identical time-dependent effects, but treatment times in the experimental-arm are biased towards early ToD (as in Figure 1, Methods). Treatment times are based on empirical data from the Hunan, China cohort^4^. In the experimental arms, half of treatment times were sampled from the first part of the day (as in figure 1, Methods). Each 3×3 block shows the inferred HRs for different combinations of early vs late ToD split times (experimental-arm treatment times were sampled below this value) and varying median OS (as followup time is fixed at 3 years, a larger median OS increases the amount of censoring). We repeat these 3×3 experiments for varying weibull shape parameter *k*(rows) and various sample size (columns). Each cell shows the median and 95% confidence interval for the HR across 1000 simulations.

**Supplementary 2.**
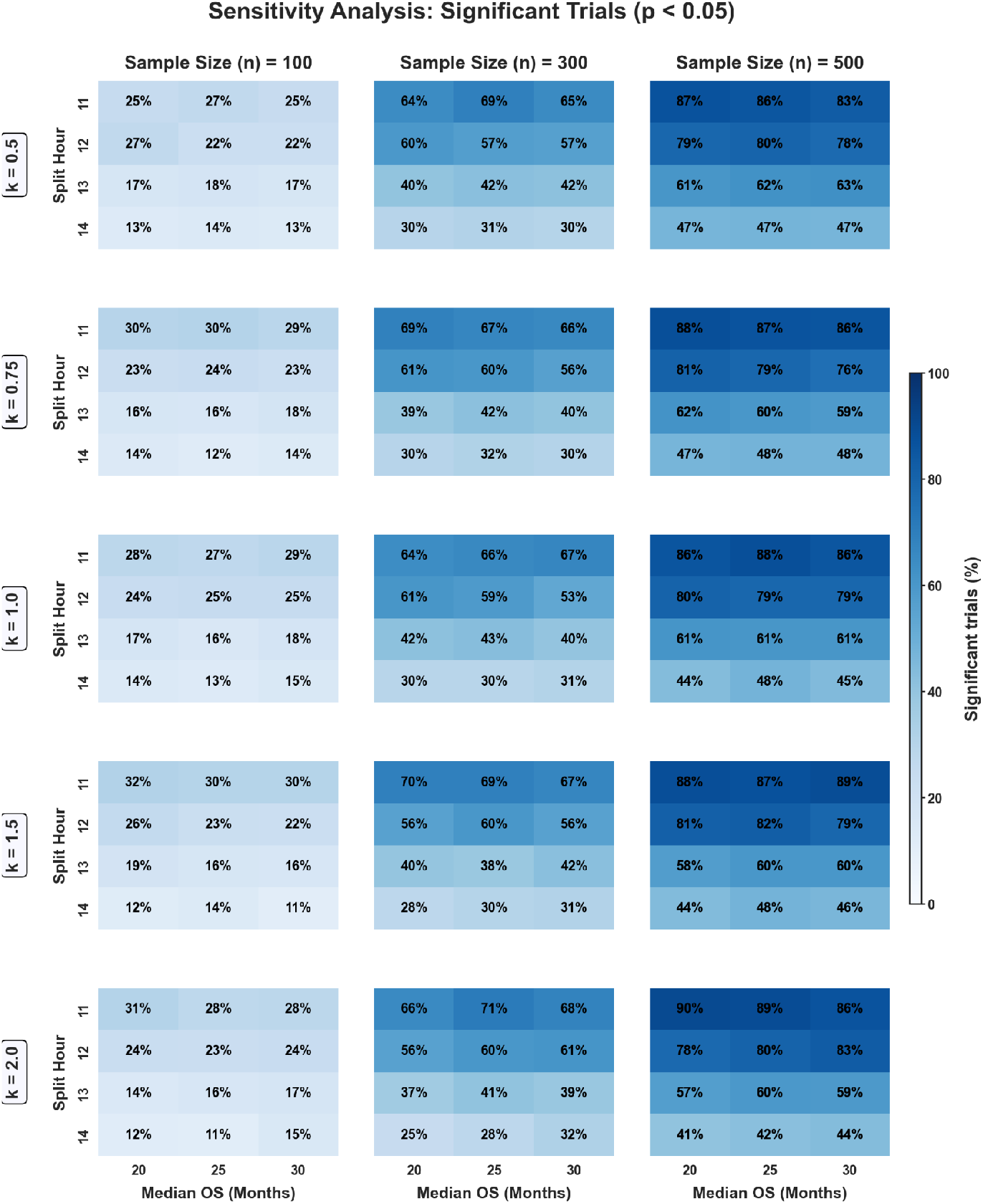
Fraction of significant trials for varying simulation parameters. Fraction of statistically significant simulations (p < 0.05), from 1000 simulated trials with 300 subjects per arm. We compared two drugs with identical time-dependent effects, where half of the treatment times in the experimental-arm are sampled from the early ToD distribution (as in Figure 1, Methods). Treatment times are based on empirical data from the Hunan, China cohort^4^. Each 3×3 block shows the fraction of significant simulations for different combinations of early vs late ToD split times (experimental-arm treatment times were sampled below this value) and varying median OS (as followup time is fixed at 3 years, a larger median OS increases the amount of censoring). We repeat these 3×3 experiments for varying weibull shape parameter *k*(rows) and various sample size (columns).

**Supplementary 3.**
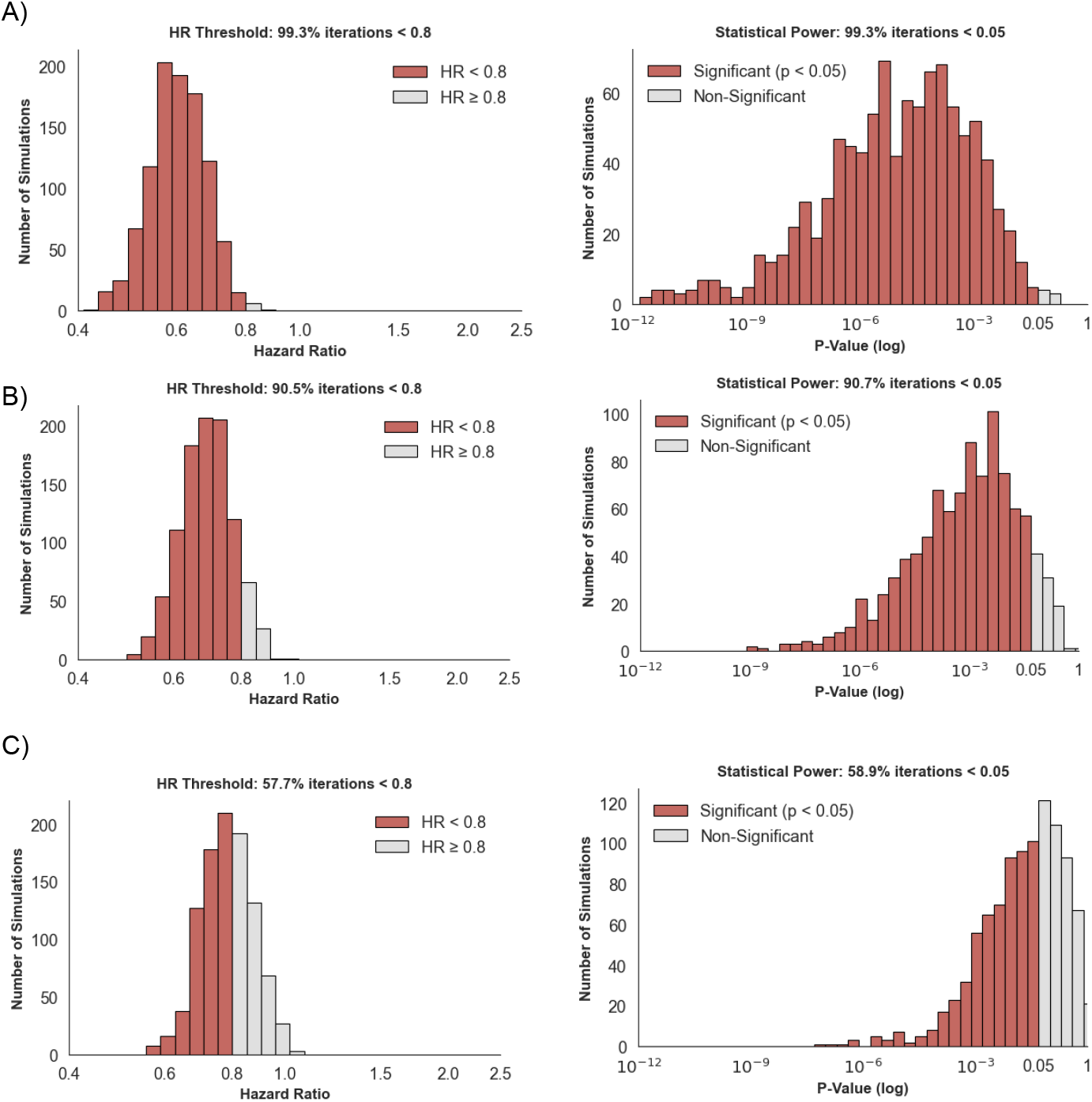
Distribution of HRs and p-values for different fractions of experimental arm patients sampled from the early ToD period. Panels A,B,C correspond with 100%, 75% and 50% of experimental arm patients sampled from the early ToD period, respectively. Each panel is based on 1000 simulations with 300 subjects per arm. Treatment time data from the Hunan, China cohort. A) Left - Median HR: 0.603 (percentiles 2.5 and 97.5: 0.4714, 0.7474). 99.3% of the experiments had a HR<0.8. Right - the median log-rank p-value was 1. 466 * 10^−5^ (percentiles 2.5 and 97.5: 1. 0849 * 10^−10^, 0.0118). 99.3% of the 1000 experiments were significant (p-value<0.05). B) Left - Median HR: 0.6933 (percentiles 2.5 and 97.5: 0.5556, 0.8555). 90.5% of the experiments had a HR<0.8. Right - The median log-rank p-value was 0.00117 (percentiles 2.5 and 97.5: 2. 5086 * 10^−7^, 0. 1703). 90.7% of the 1000 experiments were significant (p-value<0.05). C) Left - Median HR was 0.7818 (percentiles 2.5 and 97.5: 0.6315, 0.967), median p-value was 0.0278 (percentiles 2.5 and 97.5: 3. 749 * 10^−5^, 0.7518)

**Supplementary 4.**
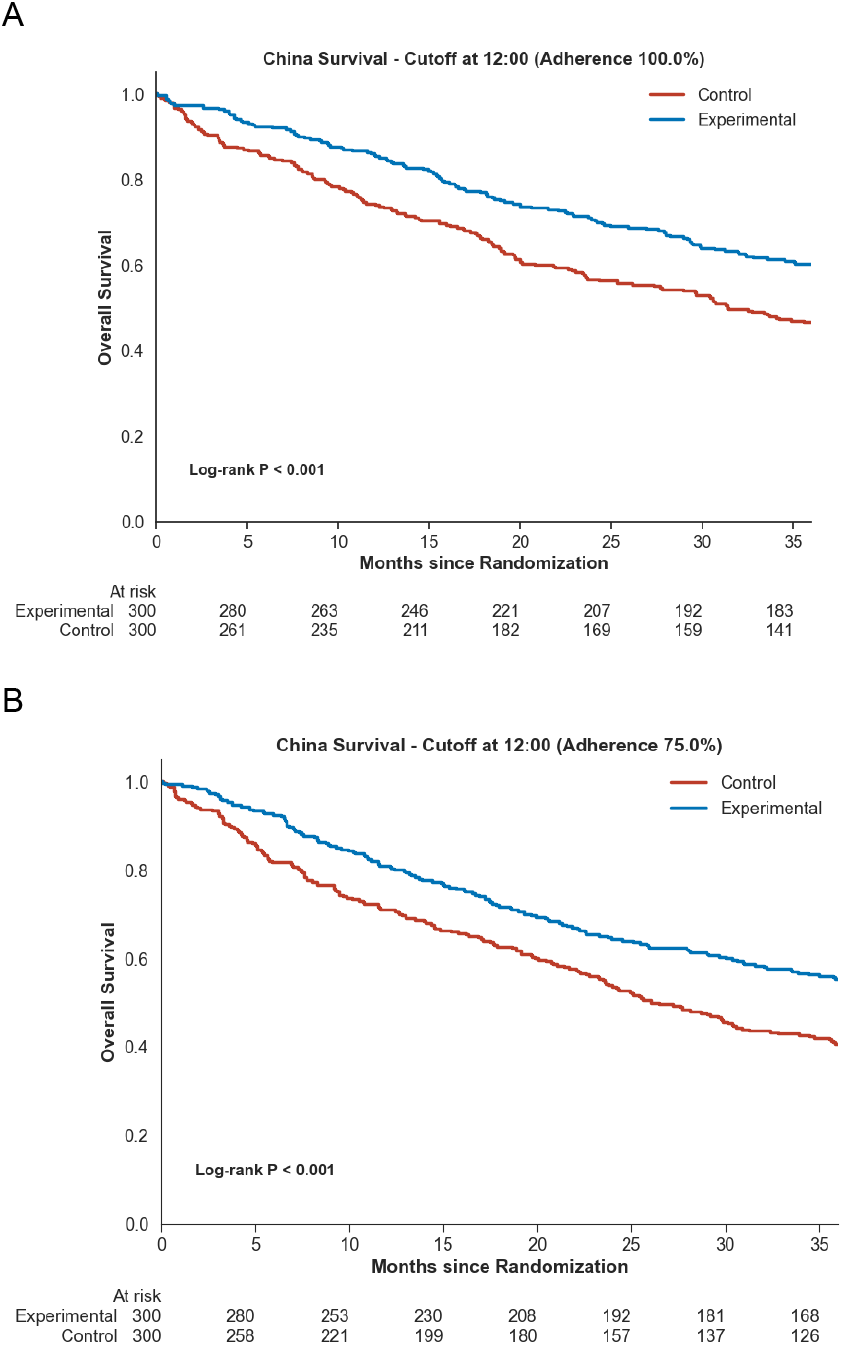
Example Kaplan Meier curves for different fractions of experimental arm patients sampled from the early ToD period. A) Example simulation where 100% of experimental arm patients were sampled from the early ToD period. Treatment time data from the Hunan, China cohort (log rank test p-value = 3. 2415 * 10^−4^). B) Example simulation where 75% of experimental arm patients are sampled from the early ToD period, respectively (log rank test p-value = 1. 8819 * 10^−4^).

**Supplementary 5.**
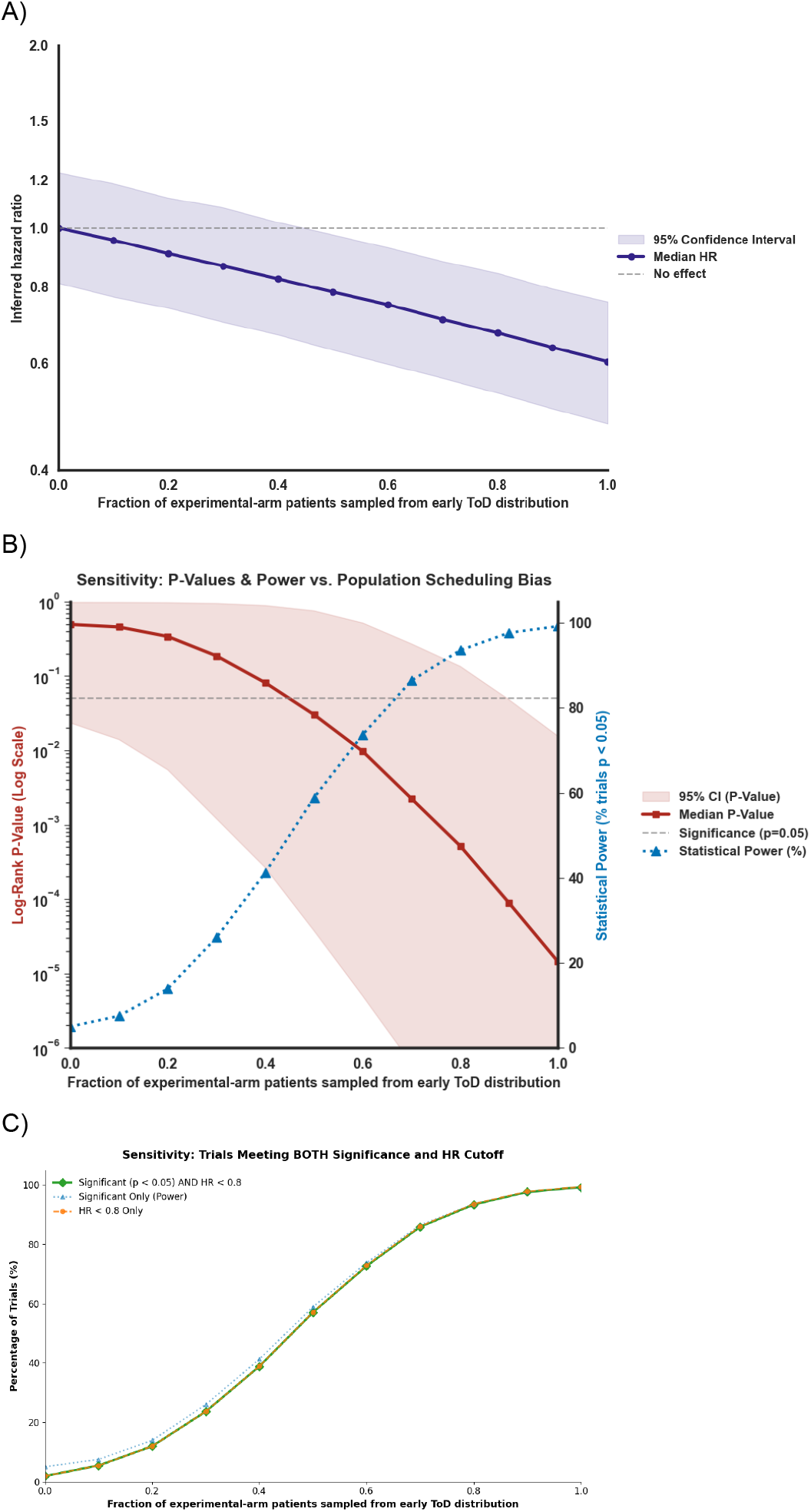
Simulations with different fractions of experimental arm subjects sampled earlier. Treatment times from the Hunan, China cohort. A) Purple line marks the median Hazard Ratio. Shaded area marks percentiles 2.5 and 97.5. B) Red line marks the median p-value (log rank test). Shaded area marks percentiles 2.5 and 97.5. Blue line marks the fraction of significant trials (“power”). C) Green line is the mean percentage of simulated trials with both significant p-value (log rank test, p<0.05) and HR<0.08. Blue dotted line shows the percentage of significant trials (p-value < 0.05) and the orange dotted line shows the percentage of trials with HR<0.08.

**Supplementary 6.**
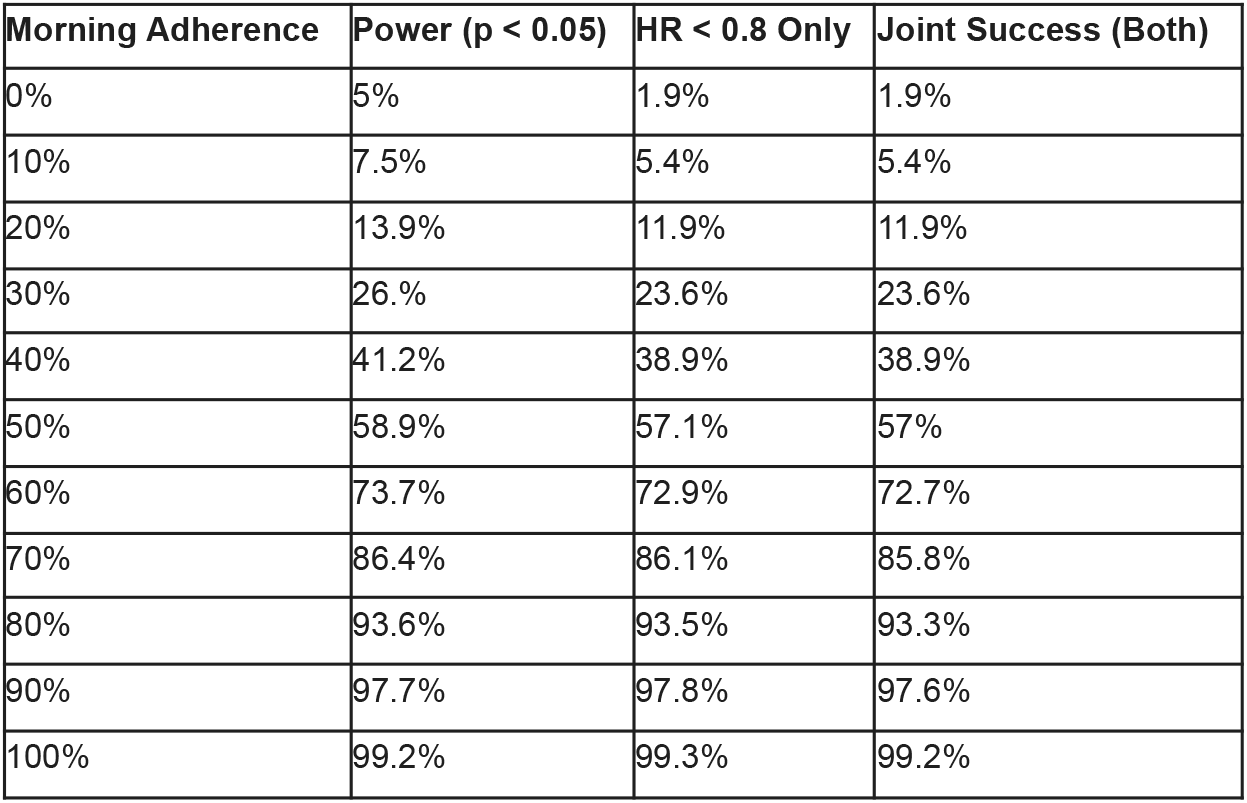
fraction of simulated trials with significant (p<0.05) and/or strong effect (HR < 0.8) for varying fractions of experiment arm patients treated earlier. Treatment times are taken from the Hunan, China cohort. Columns left to right: fraction of patients sampled from the early ToD distribution, the percentage of significant trials (log rank p-value < 0.05), the percentage of trials with HR < 0.08, the percentage of trails with both p-value<0.05 and HR<0.8.

**Supplementary 7.**
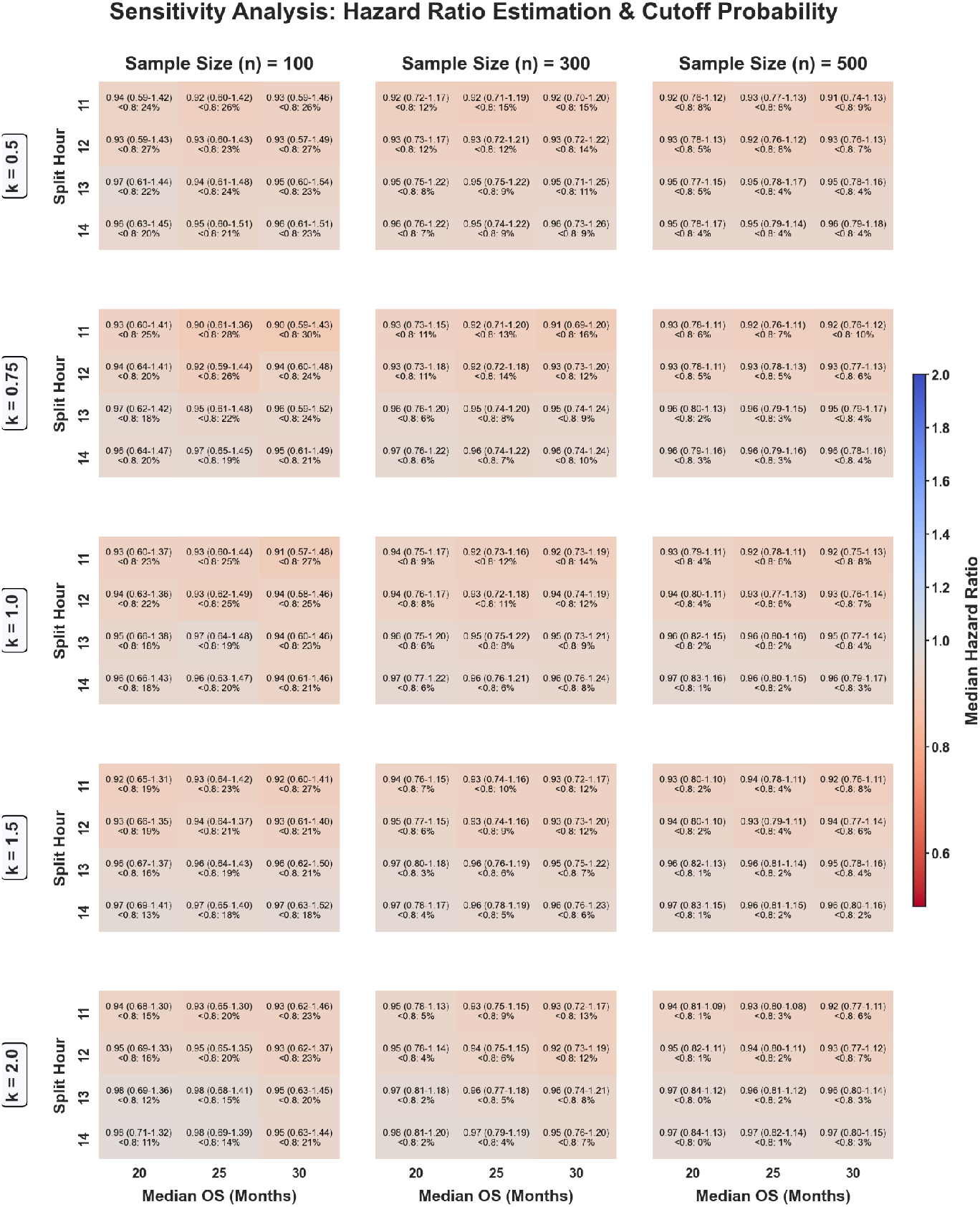
Inferred hazard ratios for varying simulation parameters. Same as supplementary 1, using treatment times from the Paris, France, cohort. In the experimental arm, half of the treatment times were sampled from the first part of the day (as in Figure 1, Methods). 300 patients per arm, 1000 simulations per setting.

**Supplementary 8.**
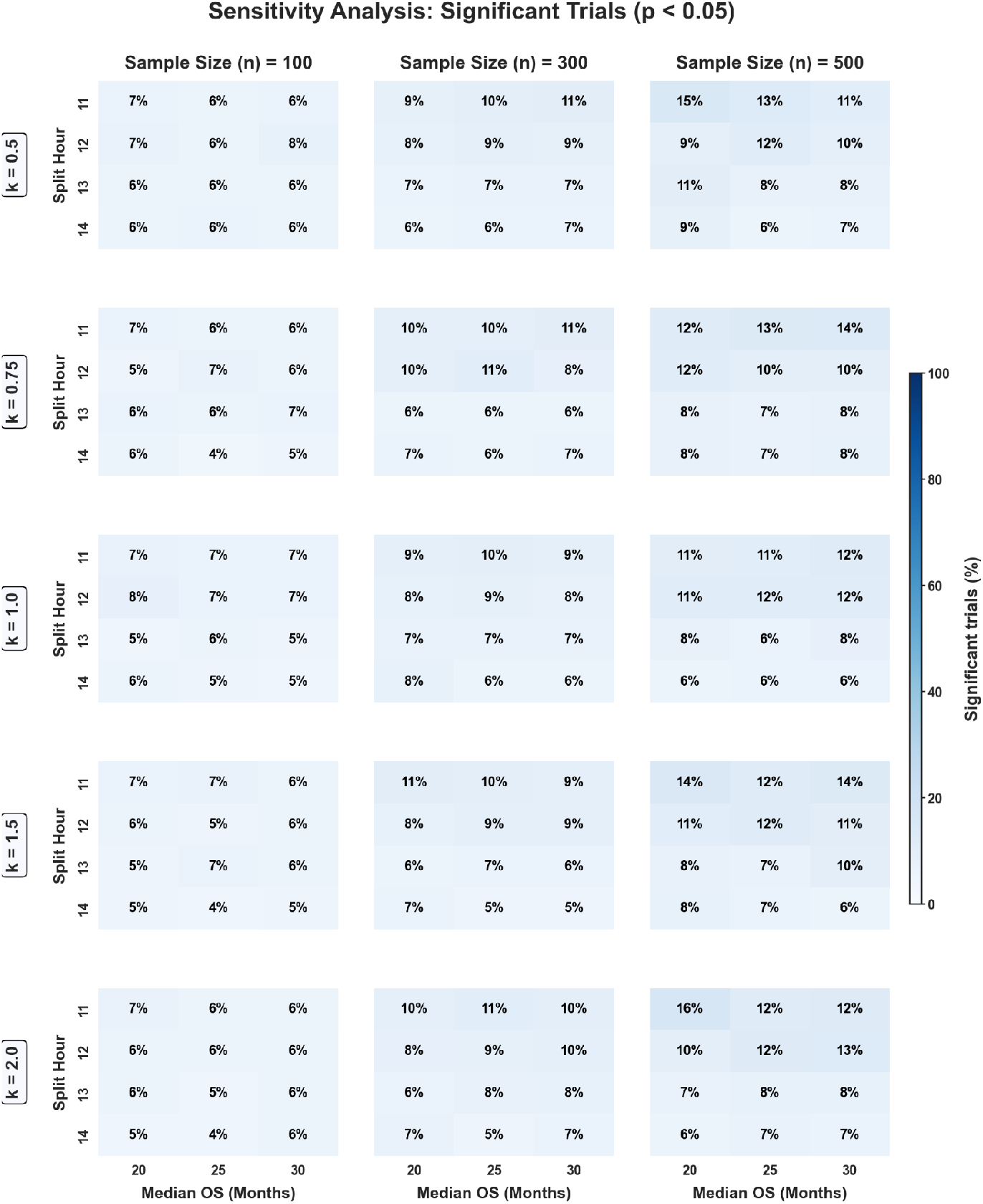
Fraction of significant trials for varying simulation parameters. Same as supplementary 2, using treatment times from the Paris, France cohort. In the experimental arm, half of treatment times are sampled from the first part of the day (as in figure 1, Methods). 300 patients per arm, 1000 simulations per setting.

**Supplementary 9.**
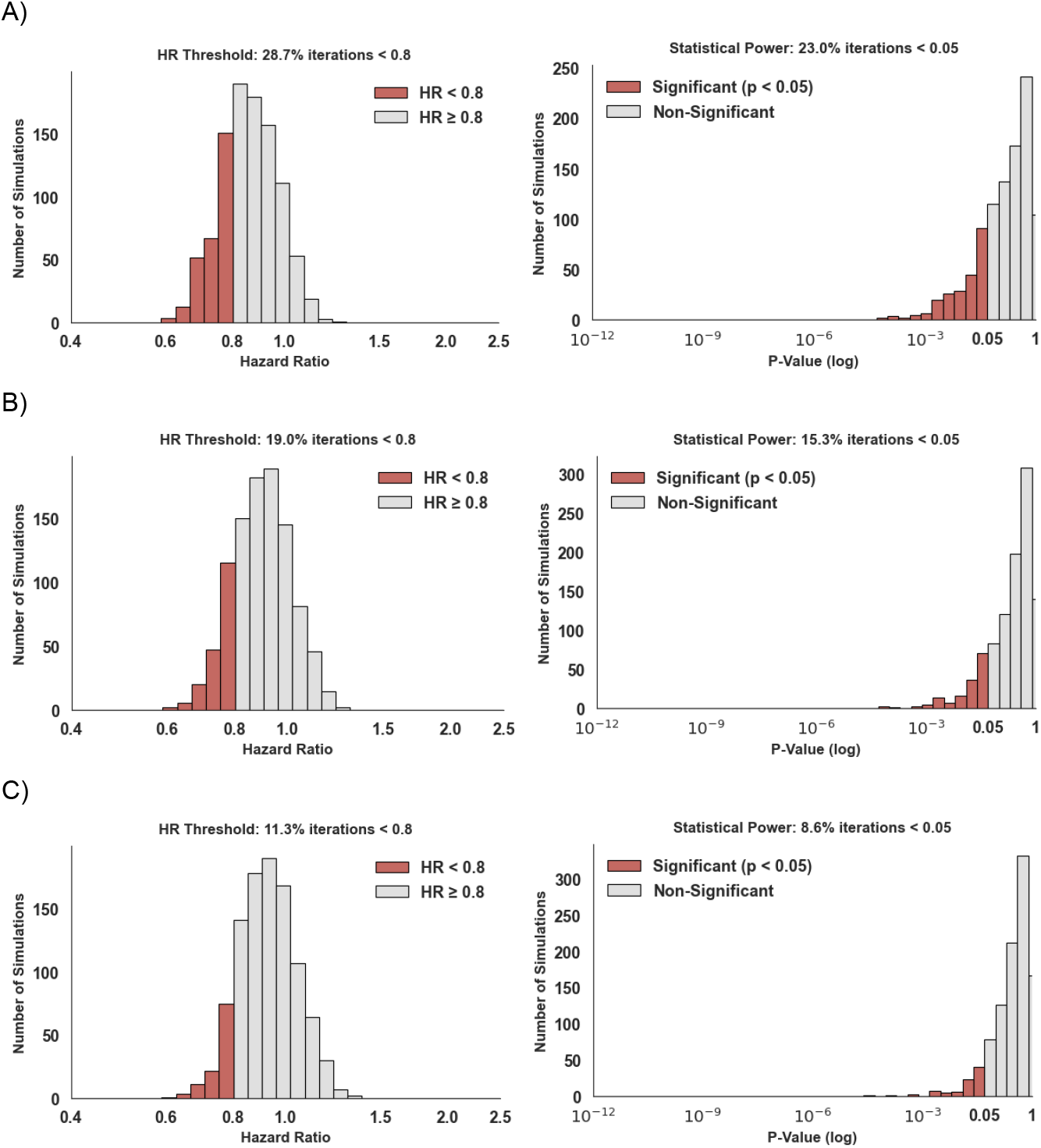
Distribution of HRs and p-values for different fractions of experimental arm patients sampled from the early ToD period. Panels A,B,C correspond with 100%, 75% and 50% of experimental arm patients sampled from the early ToD period, respectively. Treatment time data from the Paris, France cohort. Results from 1000 simulations per setting with 300 patients per arm. A) Median HR: 0.8583 (percentiles 2.5 and 97.5: 0.6803, 1.0763). 23% of the experiments had a HR<0.8, median log-rank p-value is 0.2191 (percentiles 2.5 and 97.5: 0.002, 0.9341). B) Median HR: 0.8993 (percentiles 2.5 and 97.5: 0.6972, 1.1361). 15.3% of the experiments had a HR<0.8, median log-rank p-value is 0.3428 (percentiles 2.5 and 97.5: 0.004, 0.9575). C) Median HR: 0.9238 (percentiles 2.5 and 97.5: 0.7411, 1.1728). 8.6% of the experiments had a HR<0.8, median log-rank p-value is 0.3942 (percentiles 2.5 and 97.5: 0.0148, 0.9668).

**Supplementary 10.**
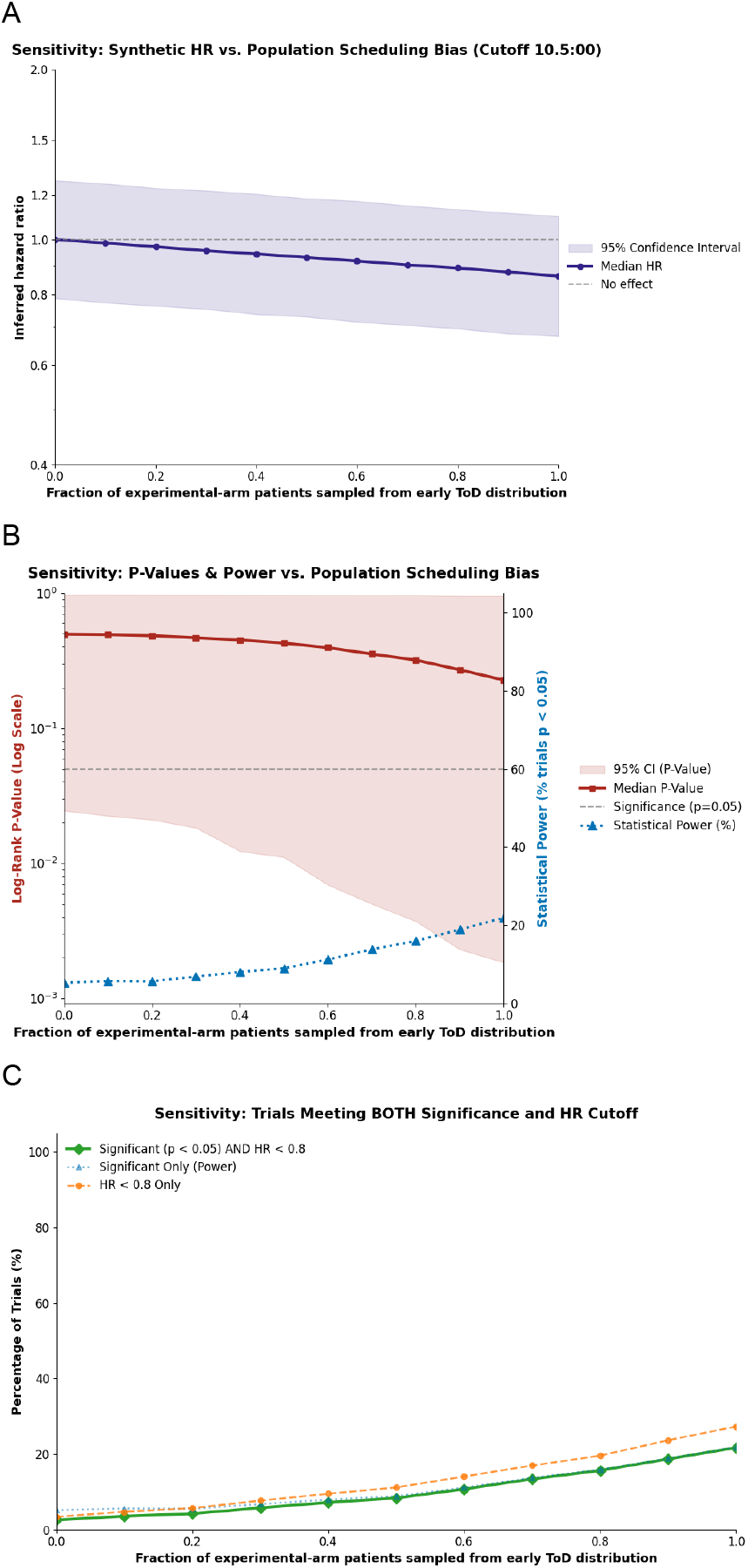
Simulations with different fractions of experimental arm subjects sampled earlier. Same as supplementary 4, using treatment times from the Paris, France cohort.

**Supplementary 11.**
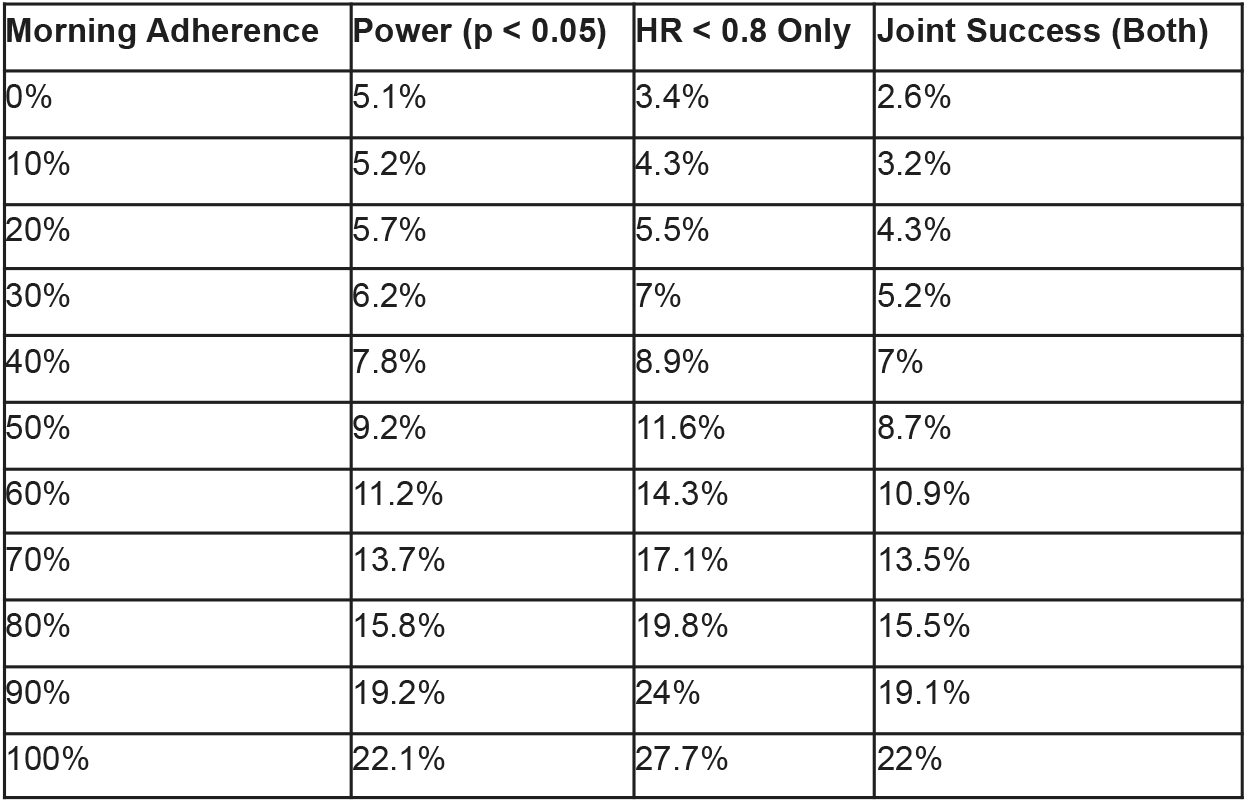
fraction of simulated trials with significant (p<0.05) and/or strong (HR < 0.8) effect for varying fractions of experiment arm patients treated earlier. Treatment times are taken from the Paris, France cohort. Columns left to right: fraction of patients sampled from the early ToD distribution, the percentage of significant trials (log rank p-value < 0.05), the percentage of trials with HR < 0.08, percentage of trails with both p-value<0.05 and HR<0.8.

**Supplementary 12.**
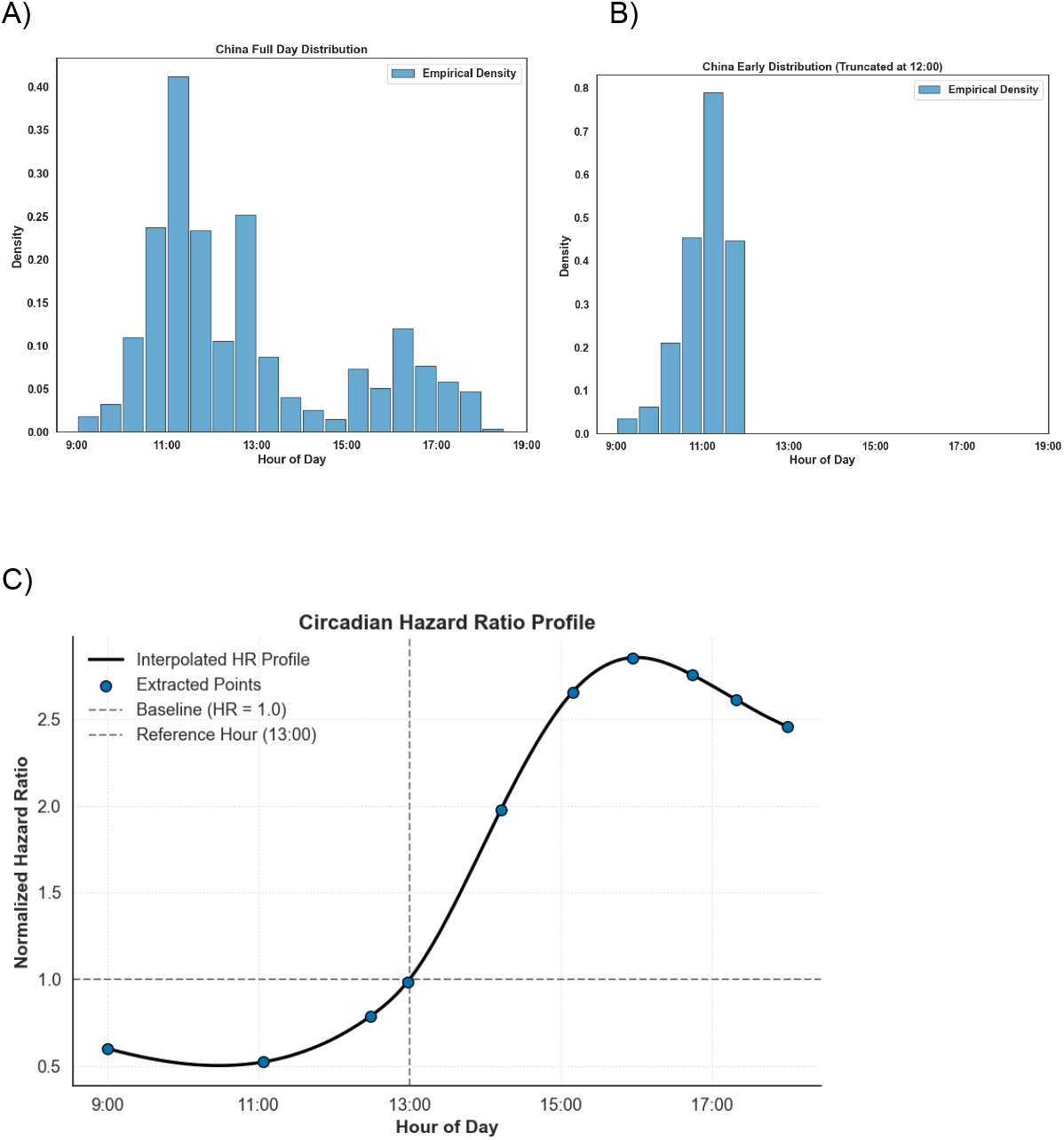
Hunan, China cohort treatment times and time-of-day dependent hazards extracted from Huang, 2025^4^. A) Distributions of patient treatment times for the whole day (China cohort, extracted from Huang, 2025 figure S1 right). B) Distribution of patient treatment times for the early ToD, namely - the empirical distribution truncated at the median treatment time. C) Hazard ratio as a function of time of day (extracted from Huang, 2025 4a). Blue points mark manually selected reference points, dark line depicts a cubic spline interpolation applied to these points. The function of hazard ratio per hour was interpolated using the extracted points (interp1d from “scipy.interpolate” version 1.13.1,with kind=‘cubic’). Hours not within the range of HR values shown in ^4^ were clipped.

**Supplementary 13.**
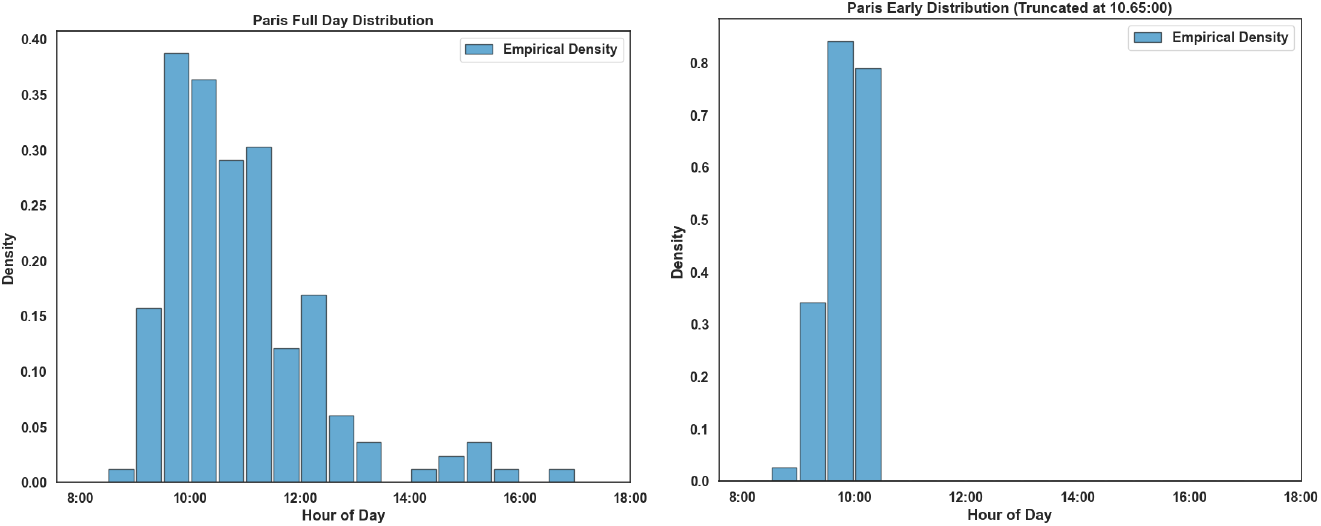
Paris, France cohort treatment times extracted from Huang, 2025^4^. A) Distributions of patient treatment times for the whole day (China cohort, extracted from Huang, 2025 figure S1 left). B) Distribution of patient treatment times for the early ToD, namely - the empirical distribution truncated at the median treatment time.

**S14.**
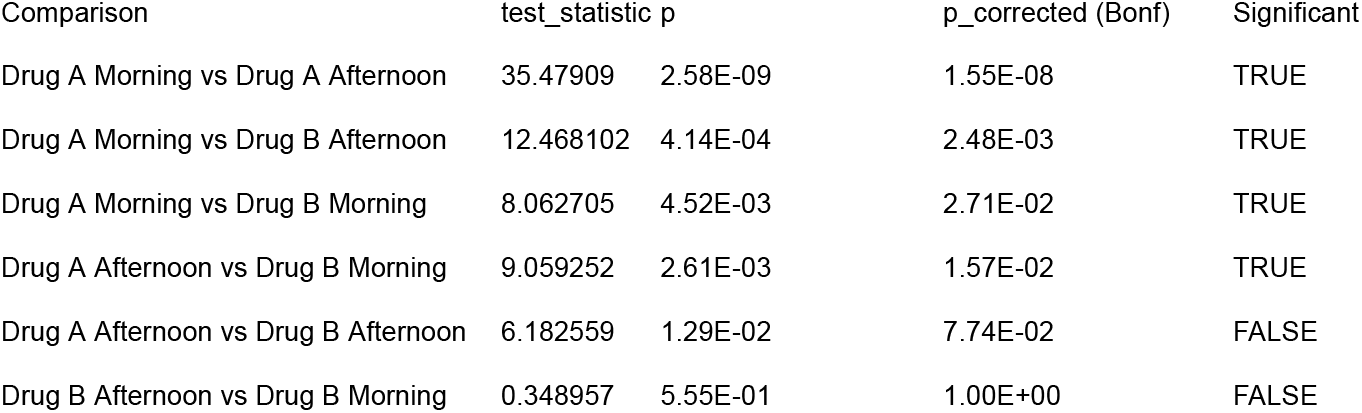
Pairwise log-rank comparisons between Drug A and B, morning and afternoon arms. Table S4 refers to the simulation in Figure 2. Each treatment arm had 300 patients (150 for drug x timing combination sub-arms). We performed pairwise log-rank tests on the simulated survival data and applied a Bonferroni correction (6 comparisons).

**S15.**
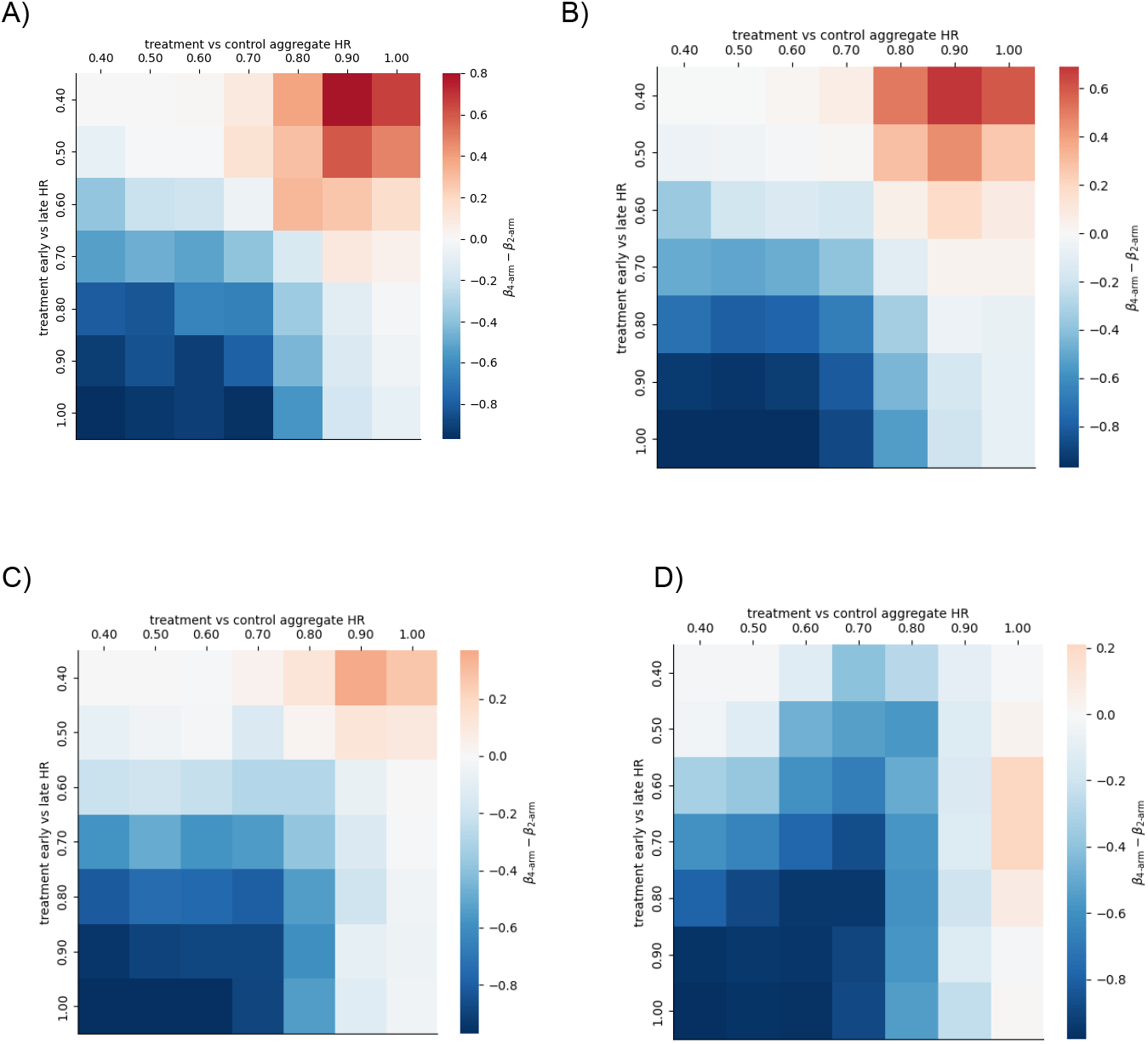
Difference in statistical power between four-arm and two-arm designs. Heatmaps display the difference in statistical power of a 2×2 design (β_4−*arm*_) and a 2-arm design (β_2−*arm*_). Regions in red mark an advantage for the 2×2 design. Each trial includes a total of n=600 subjects. This implies n=300 per arm in the 2-arm design and n=150 per arm in the 2×2 design. X-axis marks the hazard ratio between the aggregate arms (*HR*_*A,B*_) and the Y-axis marks the hazard ratio between early and late versions of the treatment arm 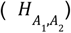. Panels A,B,C,D display analyses based on HRs of 1, 0.8, 0.6, 0.4 between early and late ToD control arms 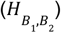, respectively. Generally, we observe that a 2×2 design may be beneficial when the aggregated main effect (A vs. B) is expected to be weaker than the within-arm temporal effect. Each setting was simulated 100 times.

## Notes

### Competing Interest Statement

The authors have declared no competing interest.

### Funding Statement

This study did not receive any funding

